# Harmonizing multisite neonatal diffusion-weighted brain MRI data for developmental neuroscience

**DOI:** 10.1101/2024.04.30.24306619

**Authors:** Alexandra F. Bonthrone, Manuel Blesa Cábez, A. David Edwards, Jo V. Hajnal, Serena J. Counsell, James P. Boardman

## Abstract

Large diffusion-weighted brain MRI (dMRI) studies in neonates are crucial for developmental neuroscience. Our aim was to investigate the utility of ComBat, and empirical Bayes tool for multisite harmonization, for removing site effects from white matter (WM) dMRI measures in healthy infants born 37-42+6 weeks from the Theirworld Edinburgh Birth Cohort (n=86) and Developing Human Connectome Project (n=287).

Skeletonized fractional anisotropy (FA), mean, axial and radial diffusivity (MD, AD, RD) maps were harmonized. The differences between voxel-wise metrics, skeleton means and histogram widths (5th-95th percentile) were assessed before and after harmonization, as well as variance associated with gestational age at birth.

Before harmonization, large cohort differences were observed in all measures. Harmonization removed all voxel-wise differences from MD maps and all metric means and histogram widths, however small voxel-wise differences (<1.5% of voxels) remained in FA, AD and RD. We detected significant relationships between GA at birth and all metrics. When comparing single site and multi-site harmonized datasets of equal sample sizes, harmonized data resulted in smaller standardized regression coefficients.

ComBat will enable unprecedented sample sizes in developmental neuroscience, offering new horizons for biomarker discovery and validation, understanding typical and atypical brain development, and assessment of neuroprotective therapies.

## 1. Introduction

Magnetic resonance imaging (MRI) studies in neonates are crucial for assessing brain development and elucidating mechanisms underlying typical and atypical neurodevelopmental outcomes. However, neurodevelopment is complex and influenced by multifactorial, often interacting neurobiological, (epi)genetic and environmental processes (Sonuga□Barke, 2023). Large, representative samples are required to disentangle these processes; this can only be achieved by using data acquired from multiple sites. Several large neonatal brain MRI cohorts exist that are now available to the research community (Boardman et al., 2020; Edwards et al., 2022; Howell et al., 2019; Soh et al., 2014); these provide new opportunities to combine datasets acquired across multiple sites. This would create unprecedented sample sizes to characterize typical population variation, delineate neurodevelopmental risk and resilience factors, identify robust and generalizable predictors of outcomes, investigate effects of (epi)genetic variation and test novel neuroprotective therapies in at-risk groups. However, there is an urgent need to identify robust strategies for combining data acquired across multiple locations.

Diffusion-weighted MRI (dMRI) measures the displacement of water molecules in tissue over time and is used to characterize microstructure in the developing brain (Pecheva et al., 2018). Mathematical models of the dMRI signal provide quantitative information which can be used to infer characteristics of the underlying tissue. The most used model is the diffusion tensor model (DTI), which provides rotationally invariant scalar metrics (fractional anisotropy [FA], and mean, axial and radial diffusivity [MD, AD, RD]) that characterize the diffusion properties of a tensor fitted to each voxel (Basser et al., 1994). DTI metrics can be calculated from standard clinical diffusion MRI sequences (Le Bihan et al., 2001) which have been widely used in neonatal dMRI research. Tensor-derived measures of white matter microstructure may be altered in neonates at risk of altered brain development such as those born preterm (Dibble et al., 2021; Vaher et al., 2022), with hypoxic ischemic encephalopathy (Dibble et al., 2020; Ward et al., 2006) and with congenital heart disease (Karmacharya et al., 2018; Miller et al., 2007; Mulkey et al., 2014). These measures are associated with both clinical and environmental risk factors such as nutrition, infection, and the perinatal stress environment (Barnett et al., 2018; Blesa et al., 2019; Boardman & Counsell, 2020; Demers et al., 2021; Lautarescu et al., 2019; Lean et al., 2022; Stoye et al., 2020; Sullivan et al., 2020l Wheater et al., 2022) and childhood neurodevelopmental outcomes in both typical and atypical populations (Dimitrova et al., 2020; K. Feng et al., 2019; Lautarescu et al., 2022; Salvan et al., 2017; Tusor et al., 2012; Ullman et al., 2015). In addition, dMRI measures have been used as markers of treatment efficacy in trials of neuroprotective therapies for neonatal populations (Azzopardi et al., 2016; Law et al., 2021; Moltu et al., 2024; O’Gorman et al., 2015; Poppe et al., 2022; Porter et al., 2010). However, dMRI-derived measures vary by both scanner and acquisition, meaning it is challenging to combine data from multiple centres.

Automated quantitative analysis of dMRI measures across the brain often requires registration to a common space, which is challenging given the large variation in brain size, shape and signal intensity across the neonatal period (Dubois et al., 2021). Methods such as tract-based spatial statistics (TBSS) (Smith et al., 2006) including its optimization for neonatal datasets (Ball et al., 2010), aim to overcome some of these challenges by non-linearly registering data, isolating a white matter ‘skeleton’ comprising of voxels with the highest FA, and projecting data from individuals onto this skeleton to ensure spatial alignment and remove contamination from non-white matter structures. Tensor-based registration algorithms such as DTI-ToolKit (DTI-TK) (Zhang et al., 2006) provide superior registration of neonatal dMRI for TBSS (Wang et al., 2011). Skeletonised DTI metrics in neonates have been analysed on a voxel-wise basis using permutation testing to identify spatially homogenous regions of alterations across the white matter skeleton (Barnett et al., 2018; Blesa et al., 2019; Mulkey et al., 2014; Porter et al., 2010).

Mean skeletonised DTI metrics can also be extracted from the whole skeleton or from a-priori hypothesised regions of the skeleton identified from a white matter atlas. However, both voxel-wise and analyses of metric averages assume homogeneity in the location and the extent of white matter alterations. Peak-width of skeletonized water diffusion metrics is a method originally developed to assess small vessel disease in adults (Baykara et al., 2016). This method measures the width of the histogram of values within the white matter skeleton (5^th^ to 95^th^ percentile). Blesa and colleagues (Blesa et al., 2020) identified correlations between MD, AD and RD histogram widths and gestational age (GA) at birth; furthermore, peak-width of skeletonised dMRI measures accurately classifies images according to GA at birth. These metrics are of particular interest for large scale studies of the neonatal brain because they characterize generalized white matter maturation and will capture changes that may be heterogenous in location and extent. In adults with small vessel disease, histogram widths were highly reproducible across MRI scanners and field strengths (Baykara et al., 2016); however, it is not known if these are reproducible across MR scanners in the newborn brain, or whether harmonization is needed to make valid inter-site comparisons.

ComBat (Johnson et al., 2007) is a data harmonization tool originally designed to remove batch effects from genomic data and is an effective harmonization tool for adult DTI data (Fortin et al., 2017). However, the effect of ComBat data harmonizationon voxel-wise, mean and histogram widths across the white matter skeleton in healthy typically developing infants has not been comprehensively assessed.

The aims of this study were to assess (i) whether FA, MD, AD and RD histogram widths are comparable across scanning sites and (ii) the utility ComBat for removing site effects from DTI measures of white matter development in healthy typically developing infants born at 37-42 weeks gestational age (‘term’). We assessed differences between sites on a voxel-wise basis and across the mean and histogram width before and after harmonization. To investigate the effect of harmonization on correlation effects, we assessed the relationship between dMRI metrics and GA at birth before and after harmonization. GA at birth was chosen as birth earlier in the term period (e.g. 37-38 weeks) is associated with altered neonatal white matter development (Broekman et al., 2014a; Gale□Grant et al., 2022; Jin et al., 2019; Ou et al., 2017a) lower neurodevelopmental scores (Hua et al., 2019; Rose et al., 2013).

## 2. Methods

The National Research Ethics Service Research Ethics Committees in West London (Developing Human Connectome Project (dHCP) 14/LO/1169) and South East Scotland (Theirworld Edinburgh Birth Cohort (TEBC) 16/SS/0154) provided ethical approvals.

In accordance with the declaration of Helsinki, informed written parental consent was obtained before MRI.

### 2.1 Participants

Preprocessed dMRI data from a subset of typically developing healthy infants at low risk of altered brain development born ≥37.0 weeks from Theirworld Edinburgh Birth Cohort (Boardman et al., 2020) and the Developing Human Connectome Project (Edwards et al., 2022) were used.

### 2.2 MRI acquisition

Infants from both cohorts underwent brain MRI during natural sleep with monitoring of pulse oximetry, electrocardiography, and temperature. All scans were supervised by a doctor or nurse trained in neonatal resuscitation and MR procedures.

Images acquired at each centre were reviewed by neuroradiologists experienced in neonatal brain imaging and all infants included in this analysis had no evidence of major incidental findings.

#### 2.2.1 Developing Human Connectome Project

Brain MRI was performed on a Philips Achieva 3 Tesla system (Best, Netherlands) situated in the neonatal intensive care unit at St. Thomas’ Hospital using a 32-channel neonatal head coil and neonatal positioning device (Hughes et al., 2017). Acoustic protection consisted of earplugs made from silicone-based putty placed in the external auditory meatus (President Putty, Coltene Whaledent, Mahwah, NJ), neonatal earmuffs (MiniMuffs; Natus Medical, Middleton, WI) and an acoustic hood placed over the infant.

dMRI was acquired with a high angular resolution diffusion multi-shell protocol designed for the neonatal brain (TR/TE 3800/90 ms, multiband acceleration factor 4, sensitivity encoding in-plane acceleration factor 1.2, in-plane resolution 1.5 × 1.5 mm, slice thickness 3 mm, 1.5 mm overlap, 300 volumes, diffusion gradient encoding: b = 0 s/mm^2^ (n = 20), b = 400 s/mm^2^ (n = 64), b = 1000 s/mm^2^ (n = 88), b = 2600 s/mm^2^ (n = 128) with 4x interleaved phase encoding)(Hutter et al., 2018).

#### 2.2.2 Theirworld Edinburgh Birth Cohort

Brain MRI was performed on a Siemens MAGNETOM Prisma 3□Tesla system (Erlangen, Germany) using a 16-channel pediatric head and neck coil. Acoustic protection consisted of flexible ear plugs and neonatal earmuffs (MiniMuffs; Natus Medical, Middleton, WI).

High angular resolution dMRI was acquired in 2 separate acquisitions to reduce the time needed to re-acquire any data lost to motion artifacts. The first acquisition consisted of 8 baseline volumes (b = 0□s/mm^2^ [b0]) and 64 volumes with b = 750□s/mm^2^, and the second multi-shell acquisition consisted of 8 b0, 3 volumes with b = 200□s/mm^2^, 6 volumes with b = 500□s/mm^2^, and 64 volumes with b = 2,500□s/mm^2^ (Acquisition parameters for both sequences: TR/TE 3500/78.0□ms; multiband acceleration factor 2, in-plane acceleration factor 2, in-plane resolution 2mm x 2mm, slice thickness 2mm).

### 2.3 dMRI preprocessing

dMRI from each cohort were preprocessed according to local procedures, described below.

#### 2.3.1 Developing Human Connectome Project

dMRI underwent parallel imaging reconstruction, denoising (Cordero-Grande et al., 2019; Veraart et al., 2016), removal of Gibbs ringing artefacts (Kellner et al., 2016), and correction for motion and image distortion using Spherical Harmonics and Radial Decomposition (Christiaens et al., 2021). The b=1000s/mm^2^ shell was extracted for further analysis.

#### 2.3.2 Theirworld Edinburgh Birth Cohort

The dMRI acquisitions underwent denoising (Veraart et al., 2016) and correction for motion and image distortion using outlier replacement and slice-to-volume registration (Andersson et al., 2003, 2016, 2017; Andersson & Sotiropoulos, 2016; Smith et al., 2004). Bias field inhomogeneity correction was performed by calculating the bias field of the mean b0 volume and applying the correction to all the volumes (Tustison et al., 2010). The first acquisition consisting of 64 directions at b=750s/mm^2^ was used for further analysis.

### 2.4 dMRI image processing

b=1000s/mm^2^ data from the dHCP and b=750s/mm^2^ were processed using the FMRIB software library (https://fsl.fmrib.ox.ac.uk/fsl/fslwiki/; FSL)(Smith et al., 2004) and DTI-TK (www.dti-tk.sourceforge.net)(Zhang et al., 2006). Diffusion tensors were calculated on a per voxel basis. Tensor images were registered to the Edinburgh Neonatal Atlas tensor template (Blesa et al., 2016, 2020) using DTI-TK. FA, MD, AD and RD maps were calculated.

A mean FA image was calculated and a mask of the mean FA skeleton was derived by perpendicular non-maximum suppression and an FA threshold of 0.15. The study-specific skeleton mask was multiplied with the custom Edinburgh neonatal atlas template skeleton mask to remove grey matter regions and fibres passing through the cerebellum, the brainstem and the subcortical grey matter (Blesa et al., 2020), based on the custom mask created by Baykara and colleagues (Baykara et al., 2016). FA, MD, AD, and RD maps for each dataset were projected onto this skeleton. For each infant, the skeleton mean and the width of the histogram (the difference between the 95th and 5th percentile voxel) for FA, MD, AD and RD were calculated.

### 2.5 Data harmonization

#### 2.5.1 ComBat

ComBat (Johnson et al., 2007) implemented in NeuroHarmonize (Pomponio et al., 2020) was used to remove the effect of site of acquisition from the skeletonized FA, MD, AD and RD maps. ComBat is an empirical Bayes method which linearly models the additive and multiplicative effects of site on feature values to minimize the variance associated with site and preserve the variance most associated with variables of interest included in the ComBat model (Fortin et al., 2017). In this analysis, the variance associated with GA at birth, GA at scan and sex were preserved.

### 2.6 Statistical analysis

All analyses except for the voxel-wise analyses were undertaken in R version 4.0.3. Differences in demographic variables between cohorts were assessed with Mann-Whitney U tests (GA at birth, GA at scan) and χ^2^ (sex).

#### 2.6.1 Voxel-wise analysis

To investigate the relationship between voxel-wise dMRI metrics, site and GA at birth before and after harmonization, voxel-wise permutation testing (10000 permutations) was performed using Randomise in FSL (http://fsl.fmrib.ox.ac.uk/fsl/fslwiki/Randomise). Significant voxels are displayed on the mean FA image and T-statistic ranges and percentage of significant voxels are reported.

##### 2.6.1.1 Differences between sites

General linear models (GLM) were used to assess the differences in dMRI metrics between dHCP and TEBC before and after harmonization with GA at birth, GA at scan and sex as covariates. Voxel-wise values before and after harmonization were extracted across the whole skeleton and mean difference plots created.

##### 2.6.1.2 Associations with GA at birth

To assess the relationship between voxel-wise dMRI metrics and GA at birth before and after harmonization, a GLM was used with GA at scan and sex as covariates.

A post-hoc analysis was undertaken to investigate the relationship between postnatal age (calculated as: GA at scan – GA at birth) and FA in regions where lower GA at birth was associated with higher FA. Mean FA values for each infant were extracted from voxels where lower FA was associated with higher GA at birth in the harmonized dataset. The association between extracted FA values and postnatal age was assessed adjusting for GA at scan and sex.

To assess the impact of data harmonization on sensitivity, we conducted voxel-wise permutation tests with five subsamples of non-harmonized data (dHCP alone, n=86 in each) and five subsamples of the full cohort of harmonized TEBC and dHCP (n=86 each). Significant voxels were extracted and the overlap of significant voxels across subsamples of un-harmonized data and the combined harmonized data.

#### 2.6.2 Mean and histogram width analyses

##### 2.6.2.1 Differences between sites

Multiple linear regressions were used to characterize the effect of site on dMRI metric means and histogram widths before and after harmonization with GA at birth, GA at scan and sex included as covariates. Standardised regression coefficients (β), standard errors and p-values are reported. The GVLMA package (Peña & Slate, 2006) assessed the validity of linear regressions, and for models where assumptions were violated, analyses were rerun with robust regression using fast-s algorithms (Lourenço et al., 2011).

##### 2.6.1.2 Associations with GA at birth

The relationship between GA at birth and dMRI metric means and histogram widths was assessed (i) before harmonization in each site separately, (ii) before harmonization without a site covariate, (iii) before harmonization with a covariate and (iv) after harmonization using the procedures described above with GA at scan and sex included as covariates.

To assess the impact of data harmonization on standardised regression coefficients, a permutation analysis was conducted with 10,000 iterative subsamples of the un-harmonized dHCP (n=86), and 10,000 iterative subsamples of the harmonized dHCP and TEBC data (n=86) with robust regressions to assess the relationship between GA at birth and dMRI metrics adjusting for GA at scan and sex. Mean standardised regression coefficients, 95% confidence intervals and standard deviations were calculated for each metric. Independent samples t-tests were used to compare standardised regression coefficients between the dHCP alone and harmonized dHCP and TEBC.

### 2.7 Data availability

Data from the dHCP is from the 3^rd^ neonatal data release (https://www.developingconnectome.org/) (Edwards et al., 2022). Requests for TEBC anonymized data will be considered under the study’s Data Access and Collaboration policy and governance process (https://www.ed.ac.uk/centre□reproductivehealth/tebc/about□tebc/for□researchers/data□access□collaboration). Scripts used to prepare data for harmonization are included as a supplement.

## 3. Results

### 3.1 Demographics

The cohorts did not differ significantly for male:female proportion or GA at birth. GA at scan was higher in TEBC compared to dHCP (Table 1).

**Table 1.** Demographics and Characteristics of dHCP and TEBC.

| Table 1. Demographics and Characteristics of dHCP and TEBC |  |  |  |
| --- | --- | --- | --- |
|  | Developing Human<br>Connectome Project<br>(n=287) | Theirworld<br>Edinburgh Birth<br>Cohort (n=86) | Differences between<br>sites |
| Gestational age at<br>birth, median (IQR) | 40.14 (39.00-40.86) | 39.71 (39.00-40.43) | p=0.076 |
| GA at scan, median<br>(IQR) | 40.86 (39.81-42.14) | 42.00 (41.29-43.00) | **p<0.001** |
| Female, n (%) | 134 (46.7) | 40 (46.5) | X<sup>2</sup>=0 p=1.0 |
| results in bold are significant |  |  |  |

### 3.2 Combat

#### 3.2.1 Voxel-wise metrics

Voxel-wise differences before and after harmonization are summarised in Figure 1. Before harmonization, 44% of FA voxels within the white matter skeleton differed between sites, with higher FA values in central white matter regions in the dHCP, and higher FA values in peripheral white matter structures in TEBC (t-statistic range −14.32-19.01). 88% of MD (t-statistic range – 11.51-27.89), 73% of AD (t-statistic range –9.95-30.90) and 86% of RD voxels (t-statistic range –12.67-23.84) within the white matter skeleton differed between sites, with higher values in TEBC compared to dHCP.

**Figure 1.**
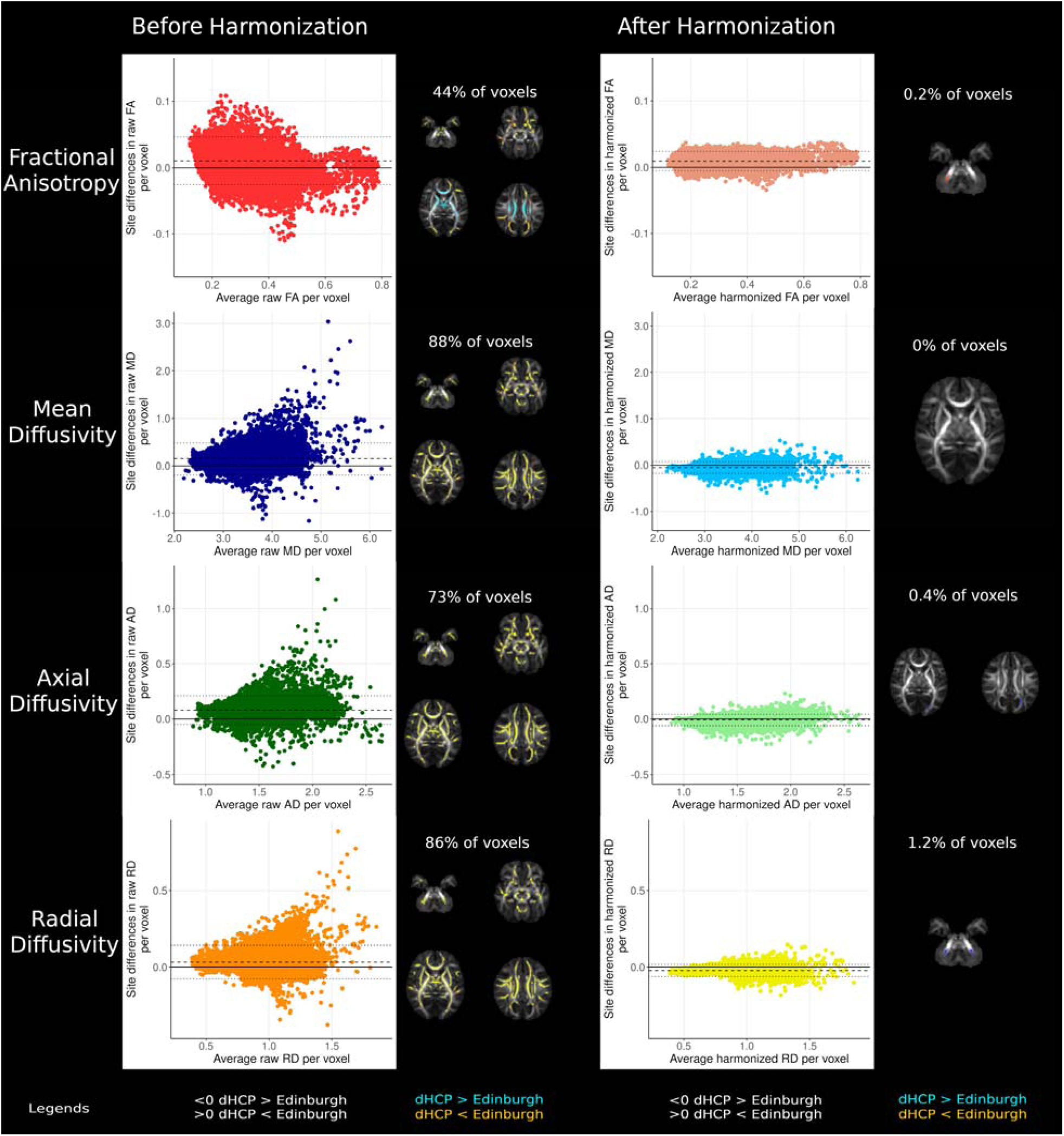
Voxel-wise differences between sites before and after harmonization. Mean difference plots with dashed lines representing mean difference and dotted lines representing 95% confidence intervals without adjusting for gestational age at birth, gestational age at scan and sex. Voxel percentages and results displayed on mean FA images represent voxels significantly different between sites after adjusting for sex, gestational age at birth and gestational age at scan.

After harmonization with ComBat, 0.2% of FA voxels were different between sites (t-statistic range −4.13-5.28), with higher values in TEBC in the right cerebellar peduncle. 0.4% of AD voxels differed between sites (t-statistic range –5.73-5.90), with higher AD in the left occipital white matter in the dHCP. 1.2% of RD voxels differed between sites (t-statistic range –5.86-4.05), with higher values in bilateral cerebellar peduncles in the dHCP. There were no significant differences in MD between sites (t-statistic range –5.82-4.94).

#### 3.2.2 Mean and histogram widths

dMRI metric means and histogram widths before and after harmonization are summarised in Table 2 and supplementary figures 1 and 2. Before harmonization, there was a significant effect of site on mean MD, AD and RD and all dMRI metric histogram widths in the white matter skeleton. After harmonization, there was no effect of site on dMRI metric means or histogram widths (Table 3).

**Table 2.** Mean and histogram widths for each site before and after harmonization.

|  |  |  |  |  |  |
| --- | --- | --- | --- | --- | --- |
| Diffusion metric | Measure, mean (SD) | Before Harmonization |  | After Harmonization |  |
|  |  | TEBC (n=86) | dHCP (n=287) | TEBC (n=86) | dHCP (n=287) |
| Fractional anisotropy | Mean | 0.265 (0.014) | 0.254 (0.019) | 0.265 (0.014) | 0.255 (0.019) |
|  | Histogram width | 0.323 (0.016) | 0.345 (0.022) | 0.343 (0.017) | 0.338 (0.022) |
| Mean Diffusivity | Mean | 3.79 (0.114) | 3.65 (0.156) | 3.64 (0.119) | 3.69 (0.155) |
|  | Histogram Width | 1.49 (0.160) | 1.57 (0.188) | 1.54 (0.168) | 1.55 (0.186) |
| Axial Diffusivity | Mean | 1.63 (0.031) | 1.55 (0.041) | 1.56 (0.031) | 1.57 (0.041) |
|  | Histogram Width | 0.702 (0.037) | 0.656 (0.048) | 0.668 (0.034) | 0.660 (0.048) |
| Radial Diffusivity | Mean | 1.08 (0.043) | 1.05 (0.059) | 1.04 (0.044) | 1.06 (0.058) |
|  | Histogram Width | 0.603 (0.058) | 0.649 (0.066) | 0.634 (0.061) | 0.637 (0.065) |

**Table 3.** Effect of site on dMRI metrics before and after harmonization.

| Table 3. Effect of site on dMRI metrics before and after harmonization |  |  |  |
| --- | --- | --- | --- |
| Diffusion metric | Measure | Effect of site before harmonization | Effect of site after harmonization |
| Fractional anisotropy | Mean | $\beta$ (SE)= -0.056<br>(0.037) $p=0.128^a$ | $\beta$ (SE)= -0.032<br>(0.037) $p=0.378^a$ |
| | Histogram width | **$\beta$ (SE)= -0.568**<br>**(0.048) $p<0.001^a$** | $\beta$ (SE)= 0.063 (0.054)<br>$p=0.243$ |
| Mean Diffusivity | Mean | **$\beta$ (SE)= -0.485**<br>**(0.040) $p<0.001$** | $\beta$ (SE)= 0.039 (0.043)<br>$p=0.360$ |
| | Histogram Width | **$\beta$ (SE)= 0.171 (0.052)**<br>**$p=0.001$** | $\beta$ (SE)= 0.014 (0.053)<br>$p=0.788$ |
| Axial Diffusivity | Mean | **$\beta$ (SE)= -0.684(0.037)**<br>**$p<0.001$** | $\beta$ (SE)= 0.048 (0.049)<br>$p=0.328$ |
| | Histogram Width | **$\beta$ (SE)= -0.376**<br>**(0.052) $p<0.001^a$** | $\beta$ (SE)=-0.006 (0.057)<br>$p=0.921^a$ |
| Radial Diffusivity | Mean | **$\beta$ (SE)= -0.370(0.040)**<br>**$p<0.001$** | $\beta$ (SE)= 0.035 (0.041)<br>$p=0.389$ |
| | Histogram Width | **$\beta$ (SE)= 0.284 (0.049)**<br>**$p<0.001$** | $\beta$ (SE)= 0.020 (0.051)<br>$p=0.692$ |
| adjusted for GA at birth, GA at scan and sex; results in bold are significant, <sup>a</sup>Robust regression |  |  |  |

#### 3.2.3 Relationship with gestational age at birth

##### 3.2.3.1 Voxel-wise metrics

Voxel-wise correlations with GA at birth before and after harmonization are summarised in Figure 2. Before harmonization, 28% of FA voxels within the white matter skeleton were associated with GA at birth. Older GA at birth was associated with higher FA values associated in the centrum semiovale, thalamus and corpus callosum and lower FA in the left posterior and retrolenticular parts of the internal capsule and bilateral cerebral peduncles (t-statistic range - 8.37-6.59). 86% of MD (t-statistic range –9.89-4.45), 88% of AD (t-statistic range –10.18-3.96) and 80% of RD voxels (t-statistic range –13.00-4.55) were negatively associated with GA at birth.

**Figure 2.**
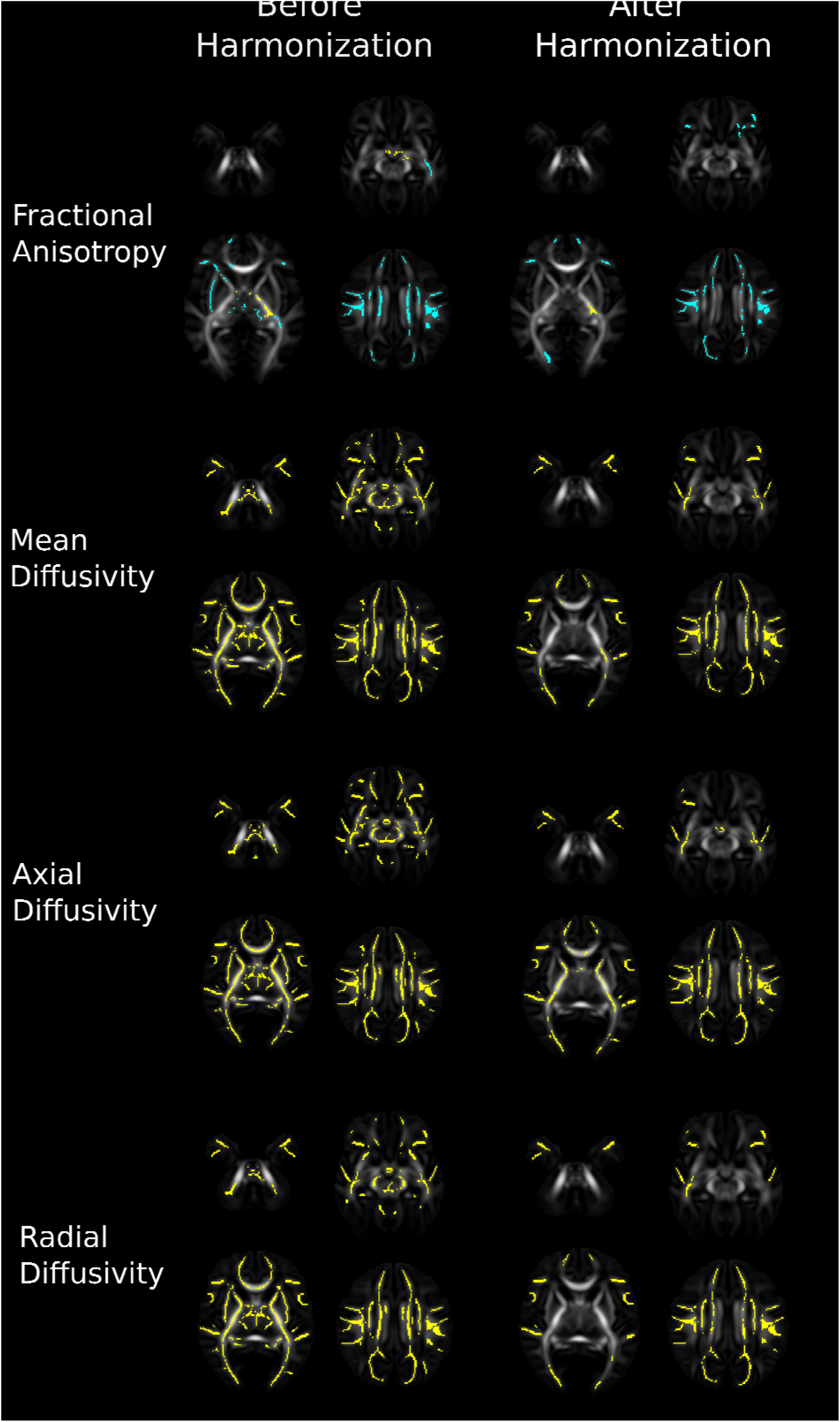
Voxel-wise correlations with GA at birth across the whole sample before and after harmonization. Results displayed on mean FA images represent voxels significantly positively (blue) and negatively (yellow) associated with gestational age at birth adjusting for sex and gestational age at scan.

After harmonization with ComBat, 22% of FA voxels were associated with GA at birth. Older GA at birth was associated with higher FA values associated in the centrum semiovale, and lower FA in the left retrolenticular part of the internal capsule (t-statistic range –6.70-6.71). Post-hoc analyses revealed FA values in the cluster within the left retrolenticular part of the internal capsule were significantly positively associated with postnatal age (β[SE]=0.358 [0.053], p<0.001), adjusting for GA at scan andsex. 46% of MD voxels (t-statistic range –10.33-4.59), 50% of AD voxels (t-statistic range –8.42-5.09), 39% of RD voxels (t-statistic range –10.48-4.92) and were negatively associated with GA at birth across the supratentorial white matter.

When iteratively assessing voxel-wise correlations with GA at birth, t-statistic ranges were similar for TEBC (n=86), five subsets of the dHCP (n=86), and five subsets of harmonized TEBC and dHCP (n=86) (Figure 3). Significant voxels identified in each iteration of dHCP and harmonized TEBC and dHCP are shown in Figure 4.

**Figure 3.**
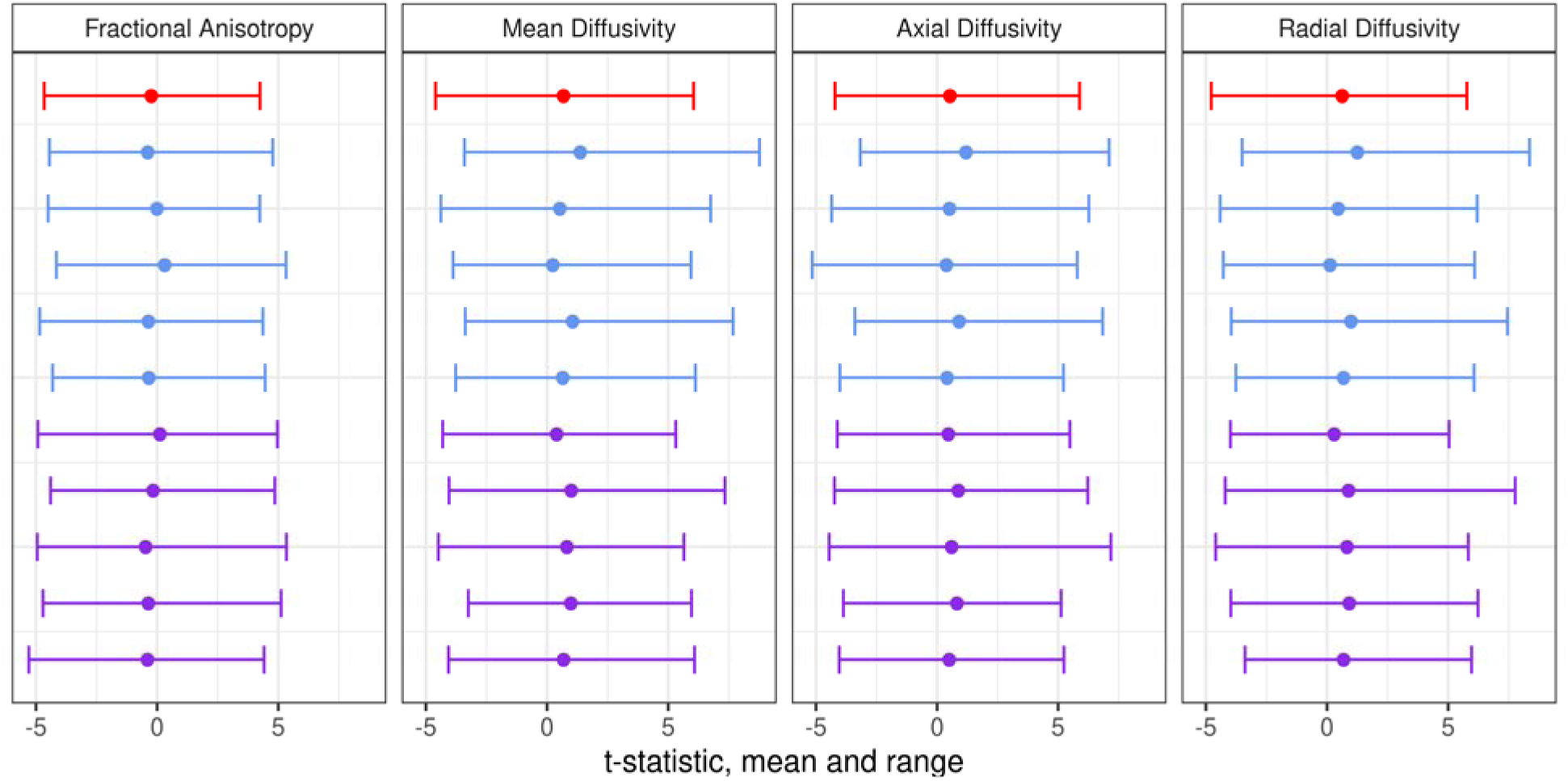
Mean and range of t-statistics for voxel-wise relationships between DTI metrics and GA at birth in Theirworld Edinburgh Birth Cohort (n=86; red), five subsamples of developing human connectome project (n=86 in each; blue) and five subsamples of combined cohort with ComBat harmonization (n=86 in each; purple).

**Figure 4.** Voxels significantly associated with GA at birth across five subsets of dHCP and harmonized dHCP and TEBC. Results displayed on mean FA images represent number of times a voxel was associated with GA at birth in each subset (0-5 times) when adjusting for sex and gestational age at scan.

##### 3.2.3.2 Mean and histogram width

Table 4 summarises the relationship between GA at birth and dMRI metric means and histogram widths in: TEBC, dHCP, the combined dataset with no correction for site, the combined dataset with site as a covariate (linear correction) and the combined dataset after ComBat harmonization. Standardised regression coefficients were similar across TEBC, dHCP and the harmonized dataset, however statistical power is higher in the combined dataset, revealing significant associations between GA at birth and mean MD, AD and RD as well as histogram widths across all metrics. Figure 5 shows the relationship between GA at birth and mean FA and histogram width in TEBC, dHCP, both datasets after ComBat harmonization and both datasets with site as a linear covariate in the regression.

**Figure 5.**
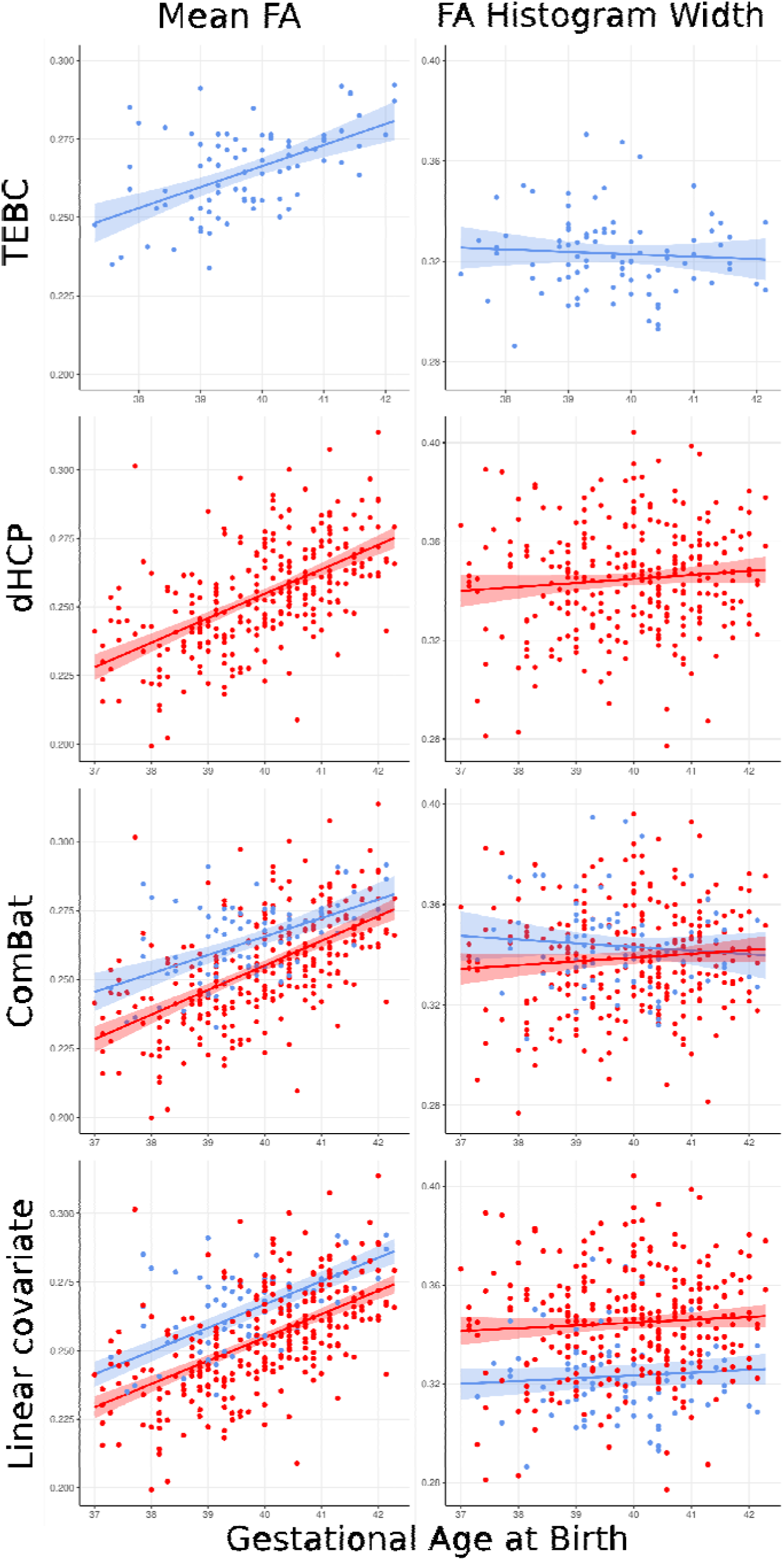
Relationship between GA at birth in Theirworld Edinburgh Birth Cohort (TEBC), developing human connectome project (dHCP) and combined cohort with ComBat harmonization and site as a linear covariate. Blue-TEBC; Red-dHCP.

**Table 4.** Effect of GA of birth on dMRI metrics before and after harmonization.

|  |  |  |  |  |  |  |
| --- | --- | --- | --- | --- | --- | --- |
| Diffusion metric | Measure | Gestational age at birth in TEBC<br>(n=86) | Gestational age at birth in dHCP<br>(n=287) | Gestational age at birth with no correction<br>(n=373) | Gestational age at birth with site as a covariate<br>(n=373) | Gestational age at birth after harmonization<br>(n=373) |
| Fractional Anisotropy | Mean | $\beta$ (SE)= 0.084<br>(0.092) p=0.363 | $\beta$ (SE)= 0.088<br>(0.057) p=0.125 | $\beta$ (SE)= 0.068<br>(0.044) p=0.127<sup>a</sup> | **$\beta$ (SE)= -0.094**<br>**(0.047) p=0.047<sup>a</sup>** | $\beta$ (SE)= 0.08<br>(0.044) p=0.072<sup>a</sup> |
| | Histogram width | **$\beta$ (SE)= -0.383**<br>**(0.130) p=0.004** | **$\beta$ (SE)= -0.248**<br>**(0.081) p=0.002** | $\beta$ (SE)= -0.001<br>(0.068) p=0.985 | **$\beta$ (SE)= -0.258**<br>**(0.062) p<0.001<sup>a</sup>** | **$\beta$ (SE)= -0.251**<br>**(0.064) p<0.001** |
| Mean Diffusivity | Mean | $\beta$ (SE)= -0.195<br>(0.109) p=0.077 | **$\beta$ (SE)= -0.243**<br>**(0.064) p<0.001** | **$\beta$ (SE)= -0.465**<br>**(0.066) p<0.001<sup>a</sup>** | **$\beta$ (SE)= -0.184**<br>**(0.015) p<0.001** | **$\beta$ (SE)= -0.212**<br>**(0.051) p<0.001** |
| | Histogram Width | $\beta$ (SE)= -0.189<br>(0.135) p=0.166 | **$\beta$ (SE)= -0.321**<br>**(0.077) p<0.001** | **$\beta$ (SE)= -0.211**<br>**(0.065) p<0.001** | **$\beta$ (SE)= -0.287**<br>**(0.066) p<0.001** | **$\beta$ (SE)= -0.291**<br>**(0.063) p<0.001** |
| Axial Diffusivity | Mean | $\beta$ (SE)= -0.245<br>(0.125) p=0.053 | **$\beta$ (SE)= -0.320**<br>**(0.072) p<0.001** | **$\beta$ (SE)= -0.569**<br>**(0.065) p<0.001<sup>a</sup>** | **$\beta$ (SE)= -0.230**<br>**(0.047) p<0.001** | **$\beta$ (SE)= -0.281**<br>**(0.058) p<0.001** |
| | Histogram Width | $\beta$ (SE)= -0.026<br>(0.136) p=0.852 | $\beta$ (SE)= -0.088<br>(0.057) p=0.125<sup>a</sup> | **$\beta$ (SE)= -0.271**<br>**(0.066) p<0.001** | $\beta$ (SE)= -0.126<br>(0.067) p=0.060<sup>a</sup> | **$\beta$ (SE)= -0.155**<br>**(0.068) p=0.023<sup>a</sup>** |
| Radial Diffusivity | Mean | $\beta$ (SE)= -0.172<br>(0.104) p=0.101 | **$\beta$ (SE)= -0.210**<br>**(0.061) p<0.001** | **$\beta$ (SE)= -0.360**<br>**(0.053) p=0.014** | **$\beta$ (SE)= -0.195**<br>**(0.051) p<0.001** | **$\beta$ (SE)= -0.183**<br>**(0.049) p<0.001** |
|  | Histogram Width | **$\beta$ (SE)= -0.294**<br>**(0.131) p=0.027** | **$\beta$ (SE)= -0.350**<br>**(0.075) p<0.001** | **$\beta$ (SE)= -0.197**<br>**(0.061) p=0.001** | **$\beta$ (SE)= -0.323**<br>**(0.062) p<0.001** | **$\beta$ (SE)= -0.338**<br>**(0.061) p<0.001** |
| results in bold are significant, <sup>a</sup>Robust regression; adjusting for GA at scan and sex |  |  |  |  |  |  |

In permutation tests, standardised regression coefficients for gestational age at birth were significantly lower in the combined harmonized data for all metrics except FA histogram width (Table 5).

**Table 5.**
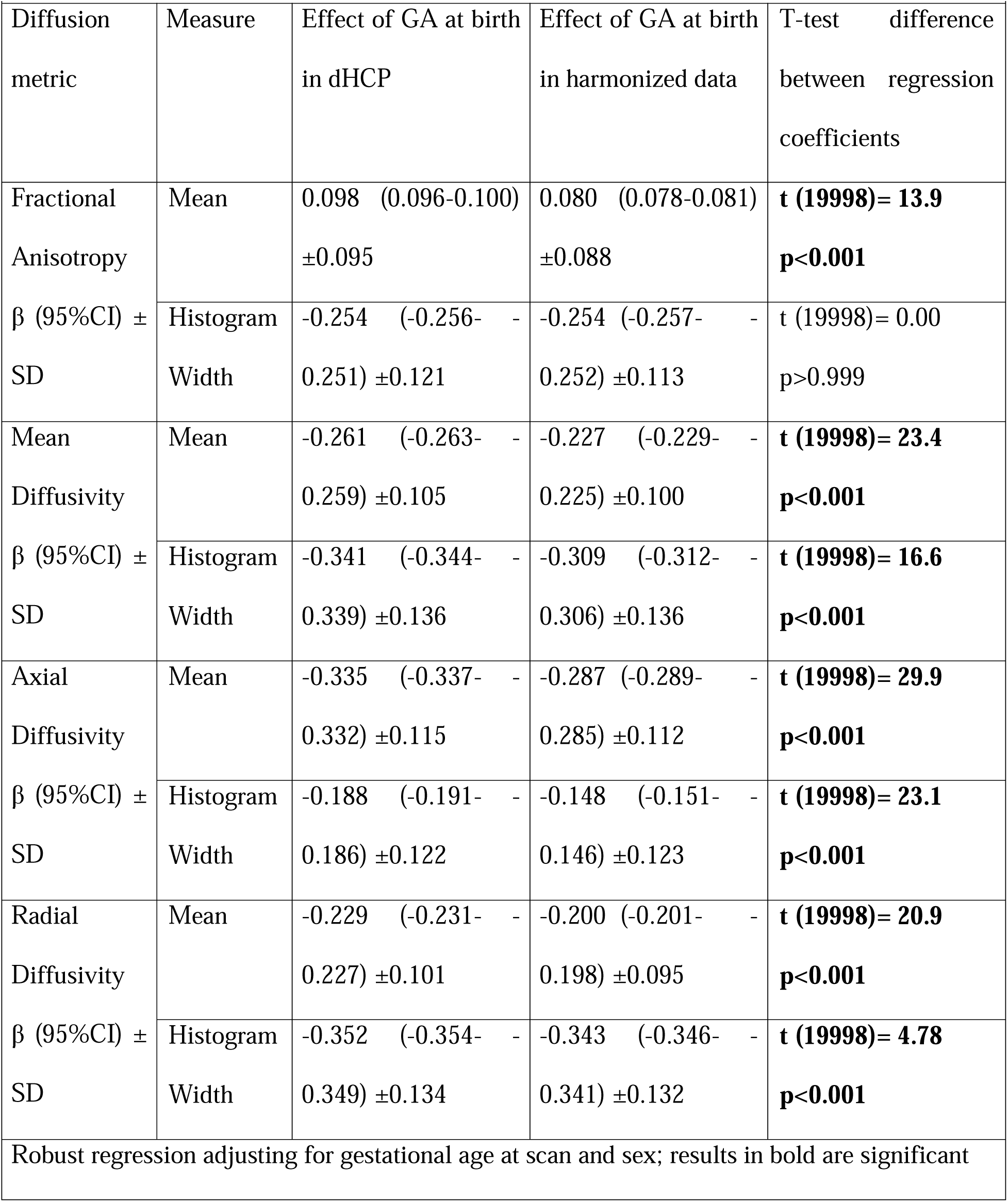
Permutation testing of the relationship between GA at birth and dMRI metric means and histogram widths.

## 4. Discussion

In this study we assessed the utility of ComBat for data harmonization across skeletonized DTI metrics in the neonatal brain. Before data harmonization, large differences in voxel-wise, mean and histogram widths were observed between two cohorts of healthy control infants who underwent neonatal brain MRI using different scanning hardware and sequence parameters. Harmonization removed all voxel-wise differences from mean diffusivity maps, however small differences (<1.5% of voxels in the white matter skeleton) remained in FA, AD and RD. Harmonization also removed significant differences in skeletonised DTI metric means and histogram widths. In the large combined harmonized dataset, we were able to detect significant relationships between GA at birth and mean and histogram widths of DTI metrics even though the study population comprised term infants only. However, when comparing single site and multi-site harmonized datasets of equal sample sizes, harmonized data resulted in smaller standardised correlation coefficients for GA at birth.

Large differences in voxel-wise DTI metrics as well as skeletonised mean (except FA) and histogram widths were observed between sites, however these were removed with ComBat harmonization. Voxel-wise mean-difference plots, and skeleton means and histogram widths, of DTI metrics after harmonization remained different between cohorts, which reflects the significantly higher GA at scan in TEBC. When adjusting for GA at scan, GA at birth and sex, there were no significant effect of site on DTI metric means or histogram widths. When combining data from multiple sites, standardizing preprocessing and quality control strategies can reduce variation between sites, however differences related to data acquisition are likely to impact extracted metrics (Galdi et al., 2024). ComBat has been used to harmonize DTI metric maps from very preterm infants imaged with different sequences on the same MRI system (Parikh et al., 2021), region of interest measures in term and preterm infants (Galdi et al., 2018), and DTI metrics along white matter tractography reconstruction in infants with congenital heart disease (Meyers et al., 2022). We add to this literature by demonstrating that ComBat successfully harmonizes voxel-wise DTI metrics as well as mean values and histogram widths from the white matter skeleton. This provides novel opportunities to retrospectively combine neonatal dMRI cohorts and overcome sample size limitations often inherent to neonatal neuroimaging research (Korom et al., 2022) and undertake large meta-analyses of neonatal neuroimaging studies. In addition, these tools may be used in clinical trials of neuroprotective ages in neonates which use dMRI an outcome, but often recruit across multiple research centers to reach adequate sample sizes.

After harmonization with ComBat and adjustment for GA at scan, GA at birth and sex, small differences in voxel-wise FA, AD, RD were found between sites. These differences may represent subtle differences between cohorts not captured in the covariates included in this analysis. Fortin and colleagues reported no white matter voxels were associated with site after ComBat harmonization of FA and MD maps in adults (Fortin et al., 2007). However, the authors used a Bonferroni correction without threshold-free cluster enhancement to control the family-wise error rate, which is more conservative than the correction used in our study and may account for the differing results. Further work with larger multi-site samples is required to investigate differences between neonatal cohorts after data harmonization.

When iteratively examining the relationship between GA at birth and mean and histogram widths of DTI metrics across 10,000 subsets of 86 babies from dHCP data and harmonized dHCP and TEBC, regression coefficients for all metrics (except FA histogram width) were significantly higher in the dHCP alone subsets. Harmonization with ComBat may therefore slightly reduce statistical power when compared to a dataset of equivalent size from one site. However, it is important to note that across the whole harmonized sample, regression coefficients were similar to TEBC (n=86), but p-values were lower reflecting the increased statistical power achieved by combining samples. Overall, harmonization improves statistical power through increased sample sizes, however some additional variance may be introduced when combining samples which is not captured by harmonization.

To our knowledge this is the largest study to assess the relationship between gestational age at birth and neonatal dMRI metrics in healthy infants born >37.0 weeks. Average mean, axial and radial diffusivity and the histogram width for all tensor metrics were associated with GA at birth. Voxel-wise TBSS analyses revealed significant associations between lower GA at birth and lower FA, and higher MD, AD and RD throughout the white matter. This may represent altered white matter development at younger GA at birth within this sample of healthy term infants. In voxel-wise TBSS studies, lower FA and higher MD, AD and RD across the brain were associated with lower GA at birth (Broekman et al., 2014; Gale□Grant et al., 2022; Jin et al., 2019; Ou et al., 2017). Interestingly, we also identified a small region of the posterior/retrolenticular part of the left internal capsule where lower GA at birth was associated with higher FA. Post-hoc analyses revealed a significant positive correlation between FA in this region and postnatal age when adjusting for sex and gestational age at scan. It is possible this reflects increased maturation of sensorimotor white matter pathways, however this requires further investigation. Our results are also in agreement with Blesa and colleagues who identified significant correlations between decreasing GA at birth and increased MD, AD and RD histogram widths in a cohort of preterm and term infants (Blesa et al., 2020). However, in contrast to Blesa and colleagues findings, a wider FA histogram in the white matter skeleton was associated with decreased GA at birth in this term-only sample. This difference is likely explained by differences in study populations as Blesa and colleagues included predominantly preterm infants. Increased mean, axial and radial diffusivity may reflect increased membrane permeability or altered oligodendrocyte proliferation (Pecheva et al., 2018), while wider FA histogram widths could represent changes to coordinate maturation patters of white matter tracts postnatally (Ouyang et al., 2019) in infants born earlier in the term period, however this requires further investigation.

It is important to note that before harmonization we identified more widespread voxel-wise associations with GA at birth across DTI metrics in regions that were significantly associated with site. Therefore, failure to appropriately remove the effect of site may introduce inflated associations in neonatal data, particularly if site is associated either with the variable of interest or necessary covariates such as GA at scan. While introducing a linear covariate may mitigate differences between sites, it’s important to note that within a GLM, the slope in each site is assumed to be equal. In our data, ComBat preserved differences in slope and data spread between sites when assessing the relationship between GA at birth and FA mean and histogram widths while linear covariates did not (as seen in Figure 4). Overall, this suggests harmonization with ComBat should be used to remove site effects rather than relying on adjusting for site with a linear covariate.

Finally, harmonizing dMRI from multiple centres to assemble large neonatal clinical cohorts has the potential to inform clinical practice. Clinical practice when treating populations in the neonatal intensive care unit can vary across centers. By harmonizing dMRI data from multiple centers to remove the effect of site but preserve the effect of clinical factors, it is possible to assess the effect of different clinical strategies on white matter development in neonates as well as disentangle the interacting effects of clinical practice and non-modifiable clinical factors. However, in the context of large multi-centre clinical neuroimaging research studies, it is necessary to ensure the definition and recording of clinical factors such as neonatal sepsis or decision-making regarding delivery management is harmonized too. Future multi-centre clinical neuroimaging research studies should engage with clinical colleagues to identify clinical variables of importance and ensure definitions are consistent between sites.

### Limitations and Future Directions

The aim of the study was to characterise the effect of ComBat on skeletonized DTI metrics in typically developing infants, therefore we did not assess the impact of data harmonization in a clinical population. Future studies should investigate the effect of ComBat on multi-centre neonatal dMRI case-control studies. In addition, further research is needed to assess the utility of novel harmonization methods such as deep learning techniques (Wada et al., 2023) for neonatal dMRI. In this study we chose to focus on harmonization of DTI metrics as these are widely used in neonatal dMRI research (Pecheva et al., 2018), and can be calculated from many conventional clinical dMRI acquisitions. However, other advanced measures of microstructure such as NODDI (Zhang et al., 2012) or fixel-based metrics (Raffelt et al., 2017) provide different insights into the developing brain and research assessing the impact of ComBat harmonization on these metrics in the neonatal brain is warranted. Finally, using dMRI to assess cortical microstructural development have reported alterations in neonates with congenital heart disease (Kelly et al., 2019) and those born prematurely (Ball et al., 2013; Galdi et al., 2024) and correlations with clinically relevant factors (Kelly et al., 2019; Sullivan et al., 2023). Future studies are required to optimize pipelines for registration and preprocessing, as well as assessing the utility of comBat for multi-centre cortical diffusion research in neonates.

### Conclusions

ComBat data harmonization removed the effect of site from skeletonised DTI metric means and histogram width in two cohorts of healthy neonates who underwent brain MRI using different scanning hardware and sequence parameters. Harmonization removed all voxel-wise differences from MD maps, however small differences remained in FA, AD and RD. In the large combined harmonized dataset, we were able to detect significant relationships between GA at birth and mean and histogram widths of DTI metrics. However, when comparing single site and multi-site harmonized datasets of equal sample sizes, harmonized data resulted in smaller standardised correlation coefficients for GA at birth. Overall, ComBat will enable the scale-up of neonatal neuroimaging studies to unprecedented sample sizes, which offers new horizons for biomarker discovery and validation, understanding upstream pathways to brain health and injury in early life, and the use of imaging for investigating the efficacy of neuroprotective therapies.

## Supporting information

scripts

## 5. Acknowledgements and Funding

This research was funded by the Harold Hyam Wingate Foundation and Medical Research Council UK (MR/V002465/1). JPB acknowledges funding from the UKRI MRC programme grant (MR/X003434/1) and Theirworld (http://www.theirworld.org/). The Developing Human Connectome Project was funded by the European Research Council under the European Union’s Seventh Framework Program (FP7/20072013)/European Research Council grant agreement no. 319456. This research was supported by core funding from the Wellcome/EPSRC Centre for Medical Engineering (WT203148/Z/16/Z) and by the National Institute for Health Research (NIHR) Biomedical Research Centre based at Guy’s and St Thomas’ NHS Foundation Trust and King’s College London. The views expressed are those of the authors and not necessarily those of the NHS, the NIHR or the Department of Health.

## Notes

### Competing Interest Statement

The authors have declared no competing interest.

