## Supplementary for "Harmonizing multisite neonatal diffusion-weighted brain MRI data for developmental neuroscience"

**Supplementary Figures**

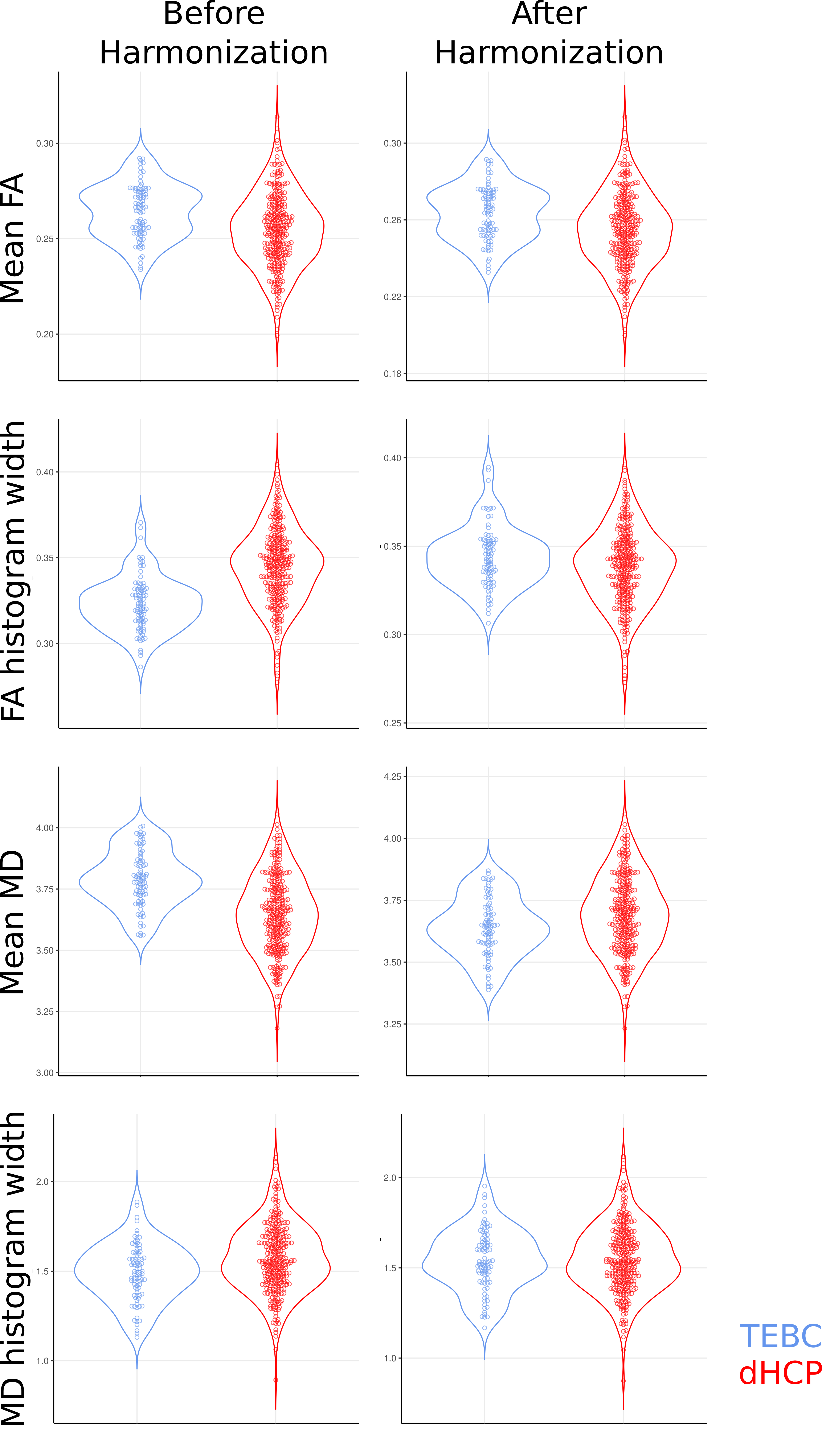

**Supplementary Figure 1.** Violin plots of mean values and histogram widths for FA and MD before and after harmonization in Theirword Edinburgh Birth Cohort (TEBC) and developing human connectome project (dHCP)

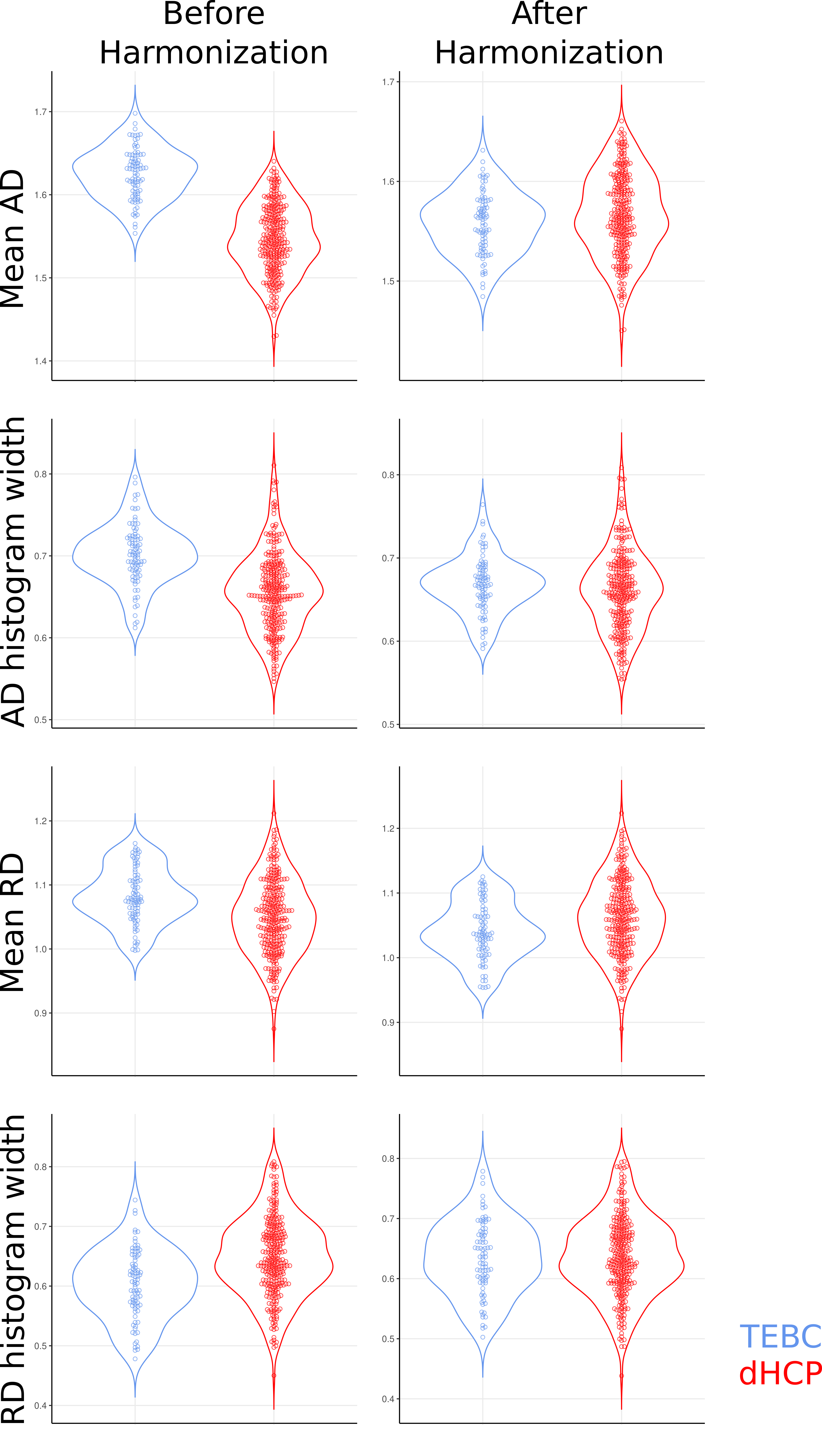

**Supplementary Figure 2.** Violin plots of mean values and histogram widths for AD and RD before and after harmonization in Theirword Edinburgh Birth Cohort (TEBC) and developing human connectome project (dHCP)

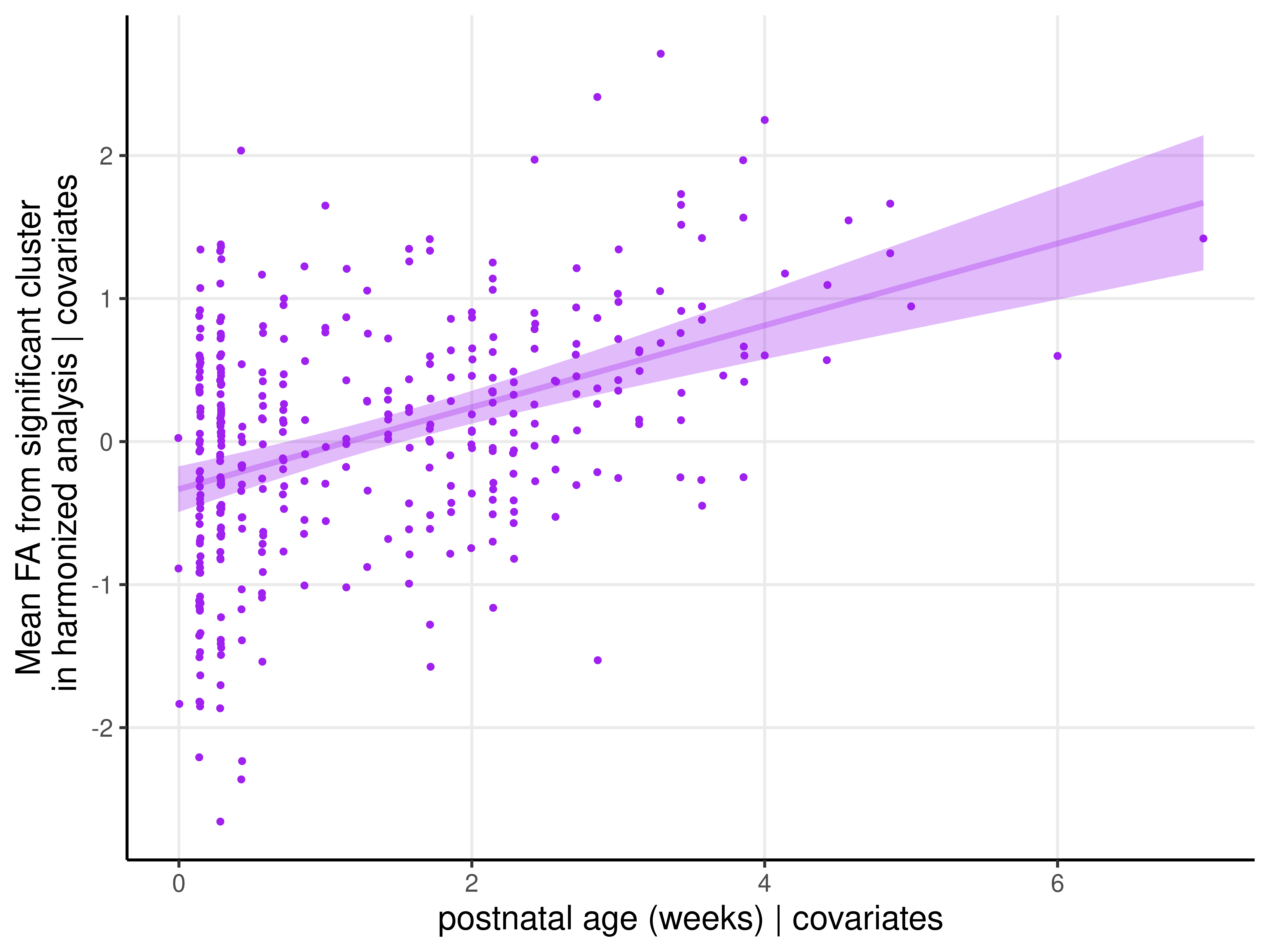

**Supplementary Figure 3.** Post-hoc analysis of relationship between FA values in the cluster within the left retrolenticular part of the internal capsule and postnatal age in weeks (β[SE]=0.358[0.053], p<0.001), adjusting for GA at scan and sex.
