## Supplementary material for "Harmonizing multisite neonatal diffusion-weighted brain MRI data for developmental neuroscience": scripts: Preprocessing_Harmonization.sh

```
#!/bin/bash

Usage() {
    echo ""
    echo "Usage"
    echo "Preprocessing_Harmonization.sh"
    echo ""
    echo "In the folder where this is gonna be excuted, there should be the aligned processed maps for each subject"
    echo "with the following convetion: subjectA_FA.nii.gz, subjectA_MD.nii.gz, etc. It can be any scalar map. In addition,"
    echo "you need to create a file called demographics.csv. This file contains information about all the covariates you"
    echo "want to include in the harmonization model. We strongly encourage to include at least gestational age at birth,"
    echo "gestational age at scan and sex. The order of the subjects in this file should match the order you can see in"
    echo "the terminal when you type ls *_FA.nii.gz. More details can be found here:"
    echo "https://github.com/rpomponio/neuroHarmonize"
    echo ""
    echo ""
    echo "Example"
    echo "Preprocessing_Harmonization.sh list_of_subjects.txt Path_of_the_ENA50"
    echo ""
    echo "The list_of_subjects.txt is a text file in the following format:"
    echo "subjectA_FA.nii.gz"
    echo "subjectB_FA.nii.gz"
    echo "..."
    echo "subjectN_FA.nii.gz"
    echo ""
    echo "The Path_of_the_ENA50 is a string with the directory where the atlas is located, for example: /home/atlas/ENA50/"
    echo ""
    echo "Script created by Alexandra Bonthrone & Manuel Blesa"
    echo ""
    exit
}
[ "$1" = "" ] && Usage

## Skeletonise all the FA maps

fslmerge -t all_FA *_FA.nii.gz

fslmaths all_FA -max 0 -Tmin -bin mean_FA_mask -odt char

fslmaths all_FA -mas mean_FA_mask all_FA

fslmaths ${2}/ENA50_FA -mas mean_FA_mask mean_FA

fslmaths mean_FA -bin mean_FA_mask

fslmaths all_FA -mas mean_FA_mask all_FA

cp ${2}/ENA50_skeleton_mask.nii.gz mean_FA_skeleton_mask.nii.gz

fslmaths mean_FA_mask -mul -1 -add 1 -add mean_FA_skeleton_mask mean_FA_skeleton_mask_dst

distancemap -i mean_FA_skeleton_mask_dst -o mean_FA_skeleton_mask_dst

tbss_skeleton -i mean_FA -p 0.15 mean_FA_skeleton_mask_dst ${2}/ENA50_LowerCingulum.nii.gz all_FA all_FA_skeletonised

## Create a folder for each metric and skeletonise them using the FA as a guide.

files=(*_MD.nii.gz)

BASENAME=` basename ${files[0]} | cut -d '_' -f 1 `

for f in ${BASENAME}*;do

mod=${f##*_}

mod=${mod%.nii.gz}

mkdir ${mod}

if [ "$mod" != "FA" ]; then

echo "Creating $mod folder"

fslmerge -t ${mod}/all_${mod} *_${mod}.nii.gz

fslmaths ${mod}/all_${mod} -mas mean_FA_mask ${mod}/all_${mod}

tbss_skeleton -i mean_FA -p 0.15 mean_FA_skeleton_mask_dst ${2}/ENA50_LowerCingulum.nii.gz all_FA ${mod}/all_${mod}_skeletonised -a ${mod}/all_${mod}

mv *_${mod}.nii.gz ${mod}/.

cp ${1} ${mod}/.

cp demographics.csv ${mod}/.

fi

done

echo "Creating FA folder"

mv *_FA.nii.gz FA/.

mv all_FA_skeletonised.nii.gz FA/.

mv mean_FA* FA/.

cp ${1} FA/.

cp demographics.csv FA/.

## Now, all the maps are in their folders and skeletonised. We need to split them and generate the files needed for the harmonization

for dir in */ ; do

mod=` basename ${dir} | cut -d '/' -f 1 `

echo "$mod"

cd ${mod}

fslsplit all_${mod}_skeletonised.nii.gz

filename="skeleton_path.csv"

header="PATH,PATH_NEW"

printf "$header\n" > $filename

j=0

while read p; do

BASENAME=` basename ${p} | cut -d '_' -f 1 `

echo ${BASENAME}

q=`printf "%0*d\n" 3 $j`

echo ${j}

cp vol0${q}.nii.gz ${j}

mv vol0${q}.nii.gz ${BASENAME}_skeletonised.nii.gz
 
printf "%s," ${PWD}/${BASENAME}_skeletonised.nii.gz >> $filename

printf "%s" ${PWD}/${BASENAME}_skeletonised_harmonized.nii.gz >> $filename

printf '\n' >> $filename

j=`expr $j + 1`

done <${1}

cd ..

done
```
